# Multi-omics profiling with untargeted proteomics for blood-based early detection of lung cancer

**DOI:** 10.1101/2024.01.03.24300798

**Authors:** Brian Koh, Manway Liu, Rebecca Almonte, Daniel Ariad, Ghristine Bundalian, Jessica Chan, Jinlyung Choi, Wan-Fang Chou, Rea Cuaresma, Esthelle Hoedt, Lexie Hopper, Yuntao Hu, Anisha Jain, Ehdieh Khaledian, Thidar Khin, Ajinkya Kokate, Joon-Yong Lee, Stephanie Leung, Chi-Hung Lin, Mark Marispini, Hoda Malekpour, Megan Mora, Nithya Mudaliar, Sara Nouri Golmaei, Hao Qian, Madhuvanthi Ramaiah, Saividya Ramaswamy, Purva Ranjan, Guanhua Shu, Peter Spiro, Benjamin Ta, Dijana Vitko, Jacob Waiss, Zachary Yanagihara, Robert Zawada, Jimmy Yi Zeng, Susan Zhang, James Yee, John E. Blume, Chinmay Belthangady, Bruce Wilcox, Philip Ma

**Affiliations:** PrognomiQ, Inc., San Mateo, CA, USA

## Abstract

Blood-based approaches to detect early-stage cancer provide an opportunity to improve survival rates for lung cancer, the most lethal cancer world-wide. Multiple approaches for blood-based cancer detection using molecular analytes derived from individual ‘omics (cell-free DNA, RNA transcripts, proteins, metabolites) have been developed and tested, generally showing significantly lower sensitivity for early-stage versus late-stage cancer. We hypothesized that an approach using multiple types of molecular analytes, including broad and untargeted coverage of proteins, could identify biomarkers that more directly reveal changes in gene expression and molecular phenotype in response to carcinogenesis to potentially improve detection of early-stage lung cancer. To that end, we designed and conducted one of the largest multi-omics, observational studies to date, enrolling 2513 case and control subjects. Multi-omics profiling detected 113,671 peptides corresponding to 8385 protein groups, 219,729 RNA transcripts, 71,756 RNA introns, and 1801 metabolites across all subject samples. We then developed a machine learning-based classifier for lung cancer detection comprising 682 of these multi-omics analytes. This multi-omics classifier demonstrated 89%, 80%, and 98-100% sensitivity for all-stage, stage I, and stage III-IV lung cancer, respectively, at 89% specificity in a validation set. The application of a multi-omics platform for discovery of blood-based disease biomarkers, including proteins and complementary molecular analytes, enables the noninvasive detection of early-stage lung cancer with the potential for downstaging at initial diagnosis and the improvement of clinical outcomes.

## Introduction

The grand hope of the National Cancer Act,^1^ passed in 1971, was that by 1976, “the final answer to cancer can be found.”^2^ After many efforts, cancer remains a leading cause of death, with lung cancer having the highest mortality in the United States (US) as well as globally.^3–5^ In 2023, US estimates of lung cancer incidence and deaths were 238,340 and 127,070, respectively, with the latter representing nearly 21% of total cancer-related deaths.^4^ The high lethality of lung cancer can be largely attributed to 53% of lung cancer cases being diagnosed as metastatic (stage IV) at initial presentation, with a correspondingly poor 5-year median overall survival of 8.2%. In contrast, for patients with localized (stage I) disease, the 5-year median overall survival improves markedly to 62.8%,^6^ although the natural history remains fatal if untreated.^7^ The disparity between these outcomes reflects the higher efficacy of the interventional armamentarium for earlier-stage disease and illustrates the value of early detection.

Lung cancers are often not diagnosed until patients develop symptoms, which are associated with late-stage disease.^8^ Thus, effective screening of asymptomatic, high-risk individuals represents a critical strategy to improve early detection, downstage initial diagnoses, and reduce mortality. The US Preventive Services Task Force (USPSTF) began endorsing screening of high-risk individuals with annual low-dose computed tomography (LDCT) scans in 2013^9^ and expanded the recommended screening population in 2021,^10^ with further expansion adopted by both the American Cancer Society (ACS) and National Comprehensive Cancer Network (NCCN).^11,12^ The implementation of annual LDCT screening has been associated with reduced mortality^13,14^ and downstaging of initial diagnoses.^15–19^ However, recent estimates of the overall screening adherence and annual adherence rates following baseline screening for eligible individuals were only 5.8%^20^ and 22.3%,^21^ respectively. These low adherence rates are influenced by various factors including patient access to LDCT and bottlenecks in clinical practice workflows as well as concerns related to increase radiation exposure^14^ and the reported false-positive rates of up to 96.4%^22^ for LDCT. These underscore the challenges of employing LDCT as a solitary screening modality in this high-risk population and highlight the magnitude of the opportunity for improvement. A peripheral blood-based biomarker test with high-performance for discriminating lung cancer, particularly at early stages, could augment current screening practices and patient access to help address this great unmet clinical need.

As cancers arise from genetic alterations,^23^ the first generation of ‘omics-based biomarker detection assays utilized genomics to survey the mutational landscape of tumor-derived DNA. Fragments of circulating tumor DNA (ctDNA) in the blood could be sequenced to detect cancer,^24,25^ albeit with concerns regarding signal-to-noise limits commensurate with tumor size^26^ and the small fraction of ctDNA relative to normal cell-free DNA (cfDNA) fragments in blood.^27^ Further advances of genomics-based cancer detection approaches have also leveraged methylation^28,29^ and fragmentation^30^ in addition to genome-wide mutational analyses.^31^ However, such approaches have a limit of detection and require a sufficient quantity of tumor-derived genetic material in blood for accurate cancer detection.^32–34^ This requirement can hinder accurate detection of early-stage cancers because the amount of ctDNA shed by small developing tumors into the blood may fall below the assay’s threshold for detection.^35,36^ The next generation of blood-based ‘omics assays applied to lung cancer detection have leveraged proteins,^37,38^ RNA,^39^ and metabolites^40,41^ with varying performance characteristics, particularly for early-stage disease.

Distinguishing true lung cancer-related biomarkers, particularly those associated with early-stage neoplastic changes, from non-cancer biomarkers related to smoking or comorbid conditions is challenging given the complexity and diversity of etiological factors contributing to lung cancer development. Thus, we posited that a multi-omics approach to both deeply and broadly interrogate the biological phenomic space of blood plasma—constituting a plurality of signal inputs from proteins, metabolites, and transcripts—would be more efficacious than individual-or dual-omics approaches to detect lung cancer, particularly at early stages. Historically, deep and large-scale untargeted surveys of the plasma proteome for biomarker discovery beyond hundreds of high abundance proteins^42,43^ has been challenging given limited throughput.^44^ However, recent developments in biomarker discovery technologies for the deep, rapid, and scalable interrogation of plasma proteins can now be applied to large-scale untargeted plasma proteomic studies^45,46^ in concert with existing discovery technologies for transcriptomics and metabolomics to enable deep multi-omics studies.

To identify a set of biomarkers that can be used to detect early-stage lung cancer with high specificity and sensitivity, we developed a multi-omics discovery approach. This approach leverages deep and untargeted exploration of the human plasma proteome with unprecedented interrogative depth and breadth at scale. Further, this approach exploits the complementarity of molecular information from additional ‘omics types (transcriptomics and metabolomics) to identify molecular signals associated with neoplastic and derivative activity. To our knowledge, this is the first time a coordinated multi-omics discovery approach has been employed at this scale in any pathology. Here, we present the development of a machine learning-based lung cancer classifier trained with multi-omics analyte data from the plasma samples of a case-control study of subjects with and without lung cancer, including those with non-malignant comorbid conditions (MOSAIC study). Evaluation of classifier performance was assessed in a separate validation set.

## Results

### Untargeted multi-analyte interrogation highlights differences in blood analytes between lung cancer and control subjects

Blood analyte data (protein, RNA, and metabolite) from subjects with and without lung cancer (N = 2513) were collected for lung cancer biomarker discovery and machine learning-based classifier building (MOSAIC study; **Figure 1**). The results reported here represent the largest known plasma multi-omics study conducted to date that uses deep, untargeted proteomics (**Figure 2**). Following quality control (QC) checks, 113,671 peptides (corresponding to 8385 protein groups) were detected in at least 1 subject, and 52,758 peptides (corresponding to 5922 protein groups) were detected in at least 25% of subjects (**Figure 2**). 83.6% (3676 proteins) of the proteins reported in the Human Plasma Proteome Project database^47^ were detected in the study subjects.

**Figure 1.**
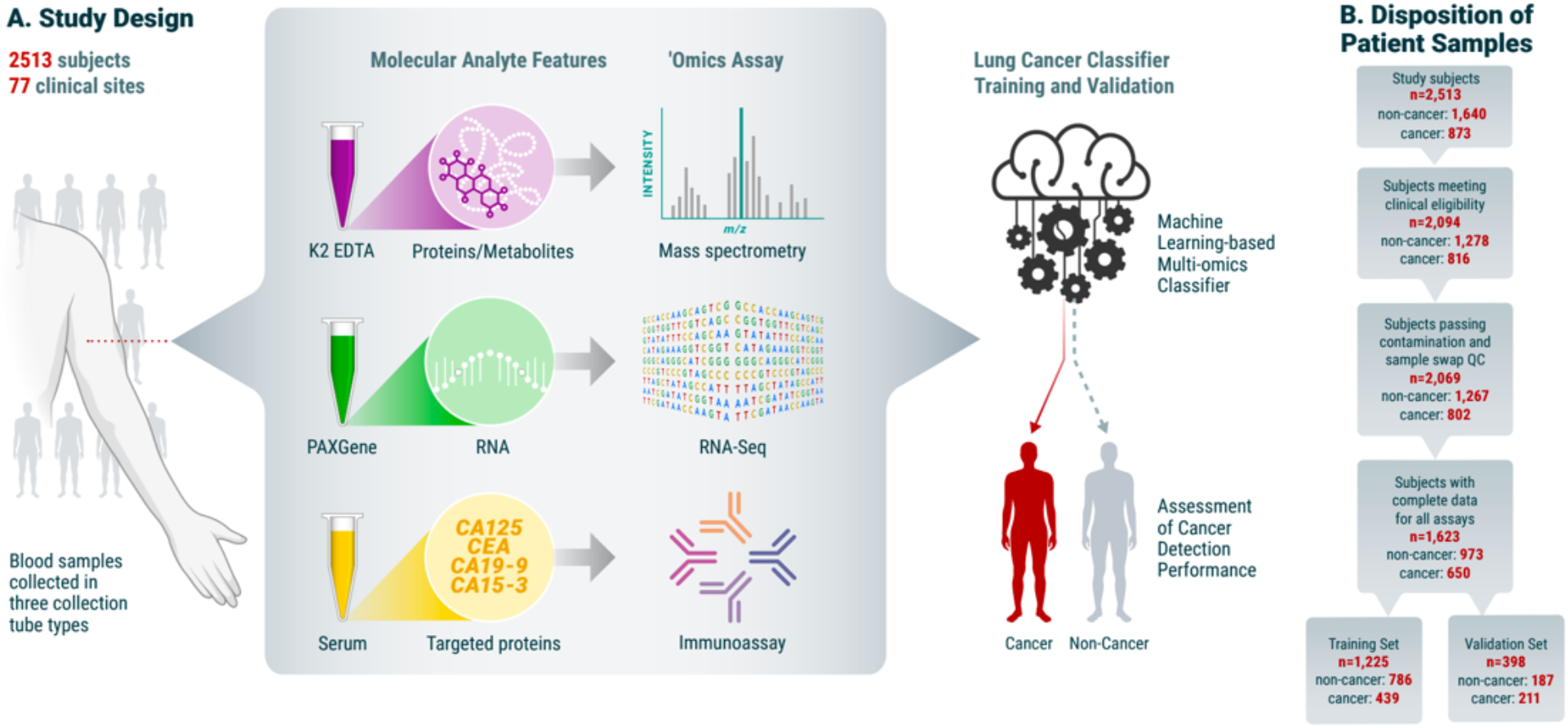
Overview of MOSAIC study. A) Subjects with and without lung cancer (N = 2,513) were enrolled in the MOSAIC study across 77 clinical sites. Three blood samples were collected per subject and used for proteomics, RNA-seq, metabolomics, and targeted immunoassays. B) Data from the ‘omics assays were then divided into training and validation sets for the development of a machine learning-based lung cancer classification model.

**Figure 2.**
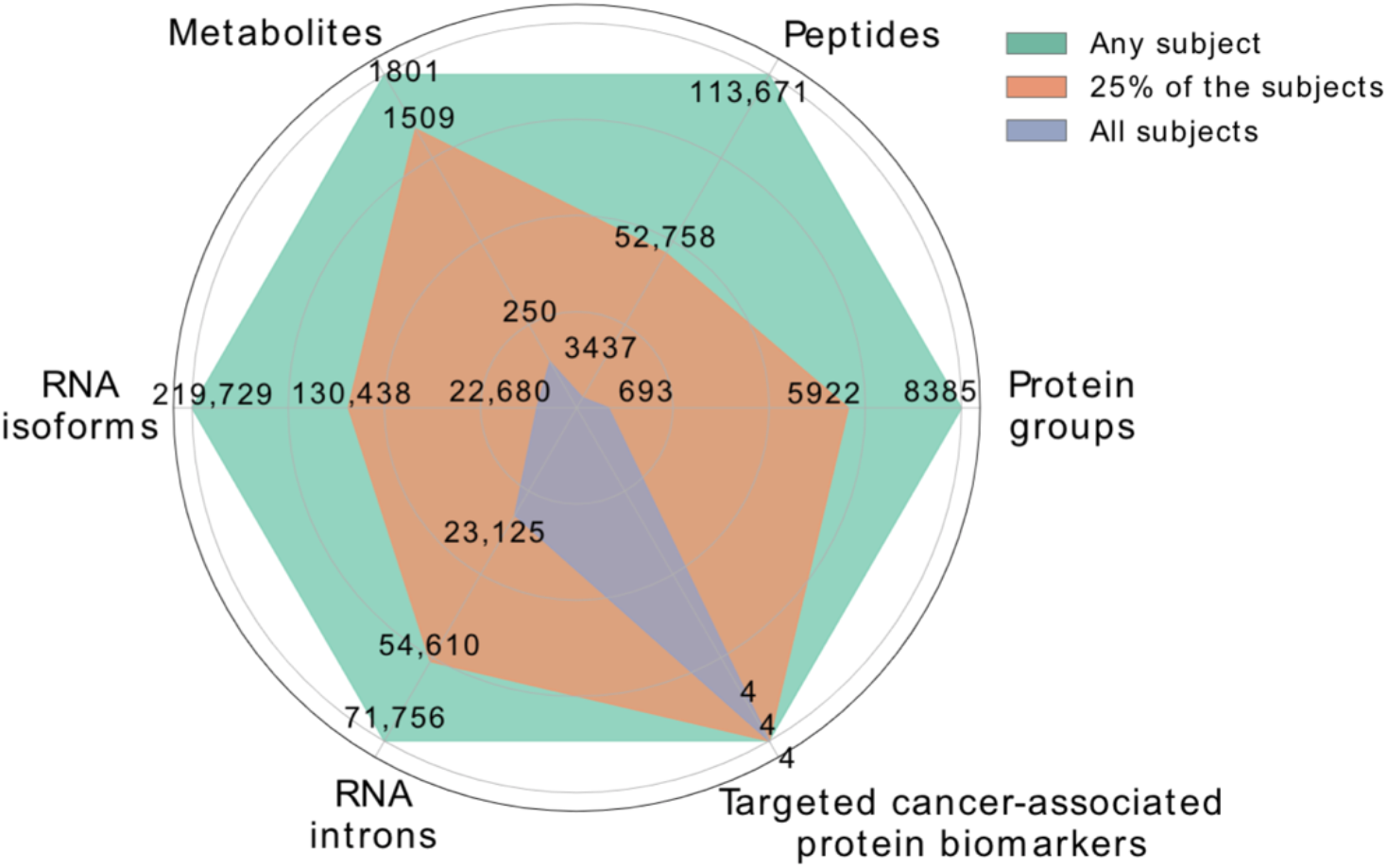
Overview of proteomics, transcriptomics, and metabolomics assays. The number of measured molecular features present in at least 1 subject (green), 25% of the subjects (orange), and all subjects (blue) for each ‘omics type.

Data were also collected from RNA-seq, metabolomics, and targeted immunoassays. Because a total RNA-seq assay was used, intronic, long non-coding, and immature RNA transcripts were also detected in the samples. In total, we detected 219,729 mRNA transcripts and 71,756 introns in at least 1 subject and 130,438 mRNA transcripts and 54,610 introns in at least 25% of the subjects. Untargeted metabolomics detected 1801 metabolites in at least 1 subject and 1509 in at least 25% of the subjects (**Figure 2**). Lastly, targeted immunoassay data focused on 4 proteins (CA125a, CA15-3, CEA, CA19-9) were collected on all subjects. Although none of these proteins are specific to a particular cancer, they are commonly used in tandem to monitor progression for various cancers.^48^

QC for enrolled subjects included verification of clinical eligibility and confirmation of data availability for all ‘omics types. Subjects passing QC were divided into 2 groups: one for training machine learning-based classifiers (training set; N = 1225) and one for validating classifier performance (validation set; N = 398) (**Figure 1B**). To begin to explore the differences in blood analytes between subjects with lung cancer and non-cancer subjects, univariate differential analysis was performed using data from the training set subjects for each ‘omics type separately. After correcting for multiple-hypothesis testing, we detected 6109 peptides, 40,171 mRNA transcripts, 9368 intronic regions, 241 metabolites, and 4 targeted proteins that were differentially abundant between the lung cancer and non-cancer cohorts (Bonferroni-corrected p-value ≤ 0.05). To understand if these differentially abundant analyte features may be identifying distinct lung cancer signals across individuals, unsupervised bi-clustering of these features was performed. Substantial heterogeneity in molecular patterns was observed within both lung cancer and non-cancer cohorts. These findings provided the rationale for supervised machine learning on multi-omics data for lung cancer classification.

### Classifiers trained on untargeted proteomics features achieved an AUC > 0.9, which was further improved by combining additional ‘omics features

We first trained a baseline classifier using only clinical variables (age, sex, and smoking status) to function as a performance comparator. The baseline classifier had an area under the receiver-operator characteristic (ROC) curve (AUC) of 0.78 (95% confidence interval [CI] 0.75-0.80) for all-stage lung cancer.

The performance of a classifier trained on only untargeted peptide features significantly outperformed this baseline model, achieving an AUC of 0.91 (95% CI 0.90-0.93) for all-stage lung cancer. To investigate if multi-omics data could further improve lung cancer classification, we trained a multi-omics classifier using analyte features from untargeted proteomics, metabolomics, RNA-seq, and the 4 immunoassayed proteins. This final multi-omics classifier had an all-stage lung cancer AUC of 0.96 (95% CI 0.96-0.97) and a stage I AUC of 0.93 (95% CI 0.92-0.95). The ROC curve for the multi-omics classifier showed that lung cancers could be detected with high sensitivity while maintaining high specificity (**Figure 3**).

**Figure 3.**
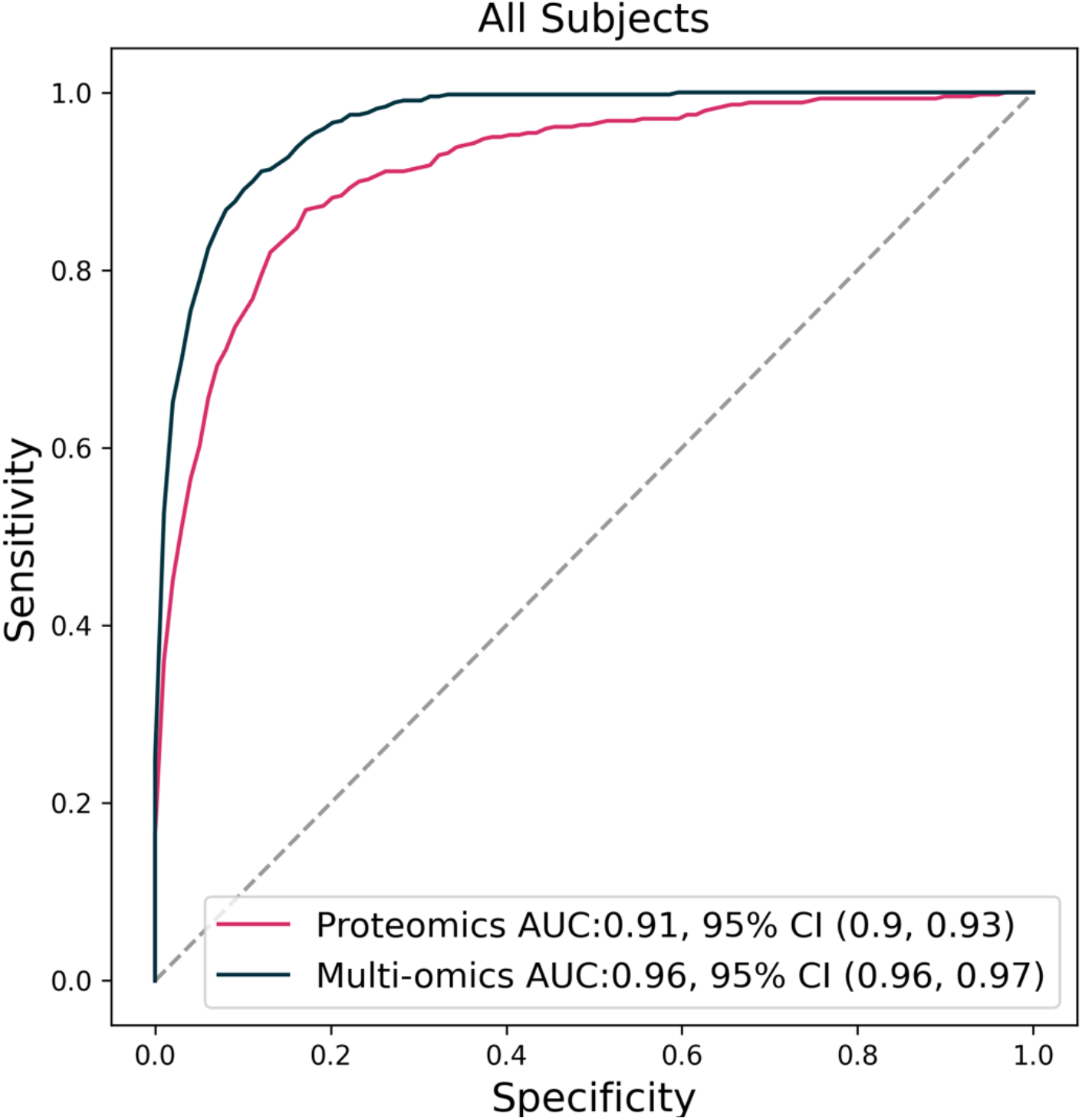
Performance of trained lung cancer classifier models. ROC curve of the multi-omics classifier (black) and untargeted proteomics classifier (pink) for all-stage lung cancers versus non-cancers.

Because the performance of the multi-omics classifier was superior to that of the baseline classifier (**Figure 3**), it was evident that the multi-omics classifier incorporated cancer-associated molecular patterns that cannot be solely attributed to age, sex, and smoking status to classify samples as cancer or non-cancer. Nonetheless, as these clinical variables were not balanced between lung cancer and non-cancer cohorts (**Table 1**), we needed to confirm that the multi-omics classifier was not fitting to these clinical variables rather than to cancer status. We evaluated the performance of the multi-omics classifier to predict sex across all subjects (AUC 0.56; 95% CI 0.53-0.59), smoking status among subjects with and without lung cancer (AUC 0.64; 95% CI 0.53-0.74 and AUC 0.65; 95% CI 0.61-0.69, respectively), and binarized age (above or below the median value of 67 years) among subjects with and without lung cancer (AUC 0.53; 95% CI 0.47-0.60 and AUC 0.69; 95% CI 0.66-0.73, respectively). These results were significantly better than random (AUC = 0.5), but much worse than what was seen for lung cancer classification, reinforcing that the multi-omics classifier is predictive of cancer status specifically. Since the clinical variables were not used as inputs during classifier training, we further surmised that these non-random results may reflect molecular signatures of sex, age, and smoking linked to cancer, as all 3 clinical variables are themselves risk factors of disease.

**Table 1.**
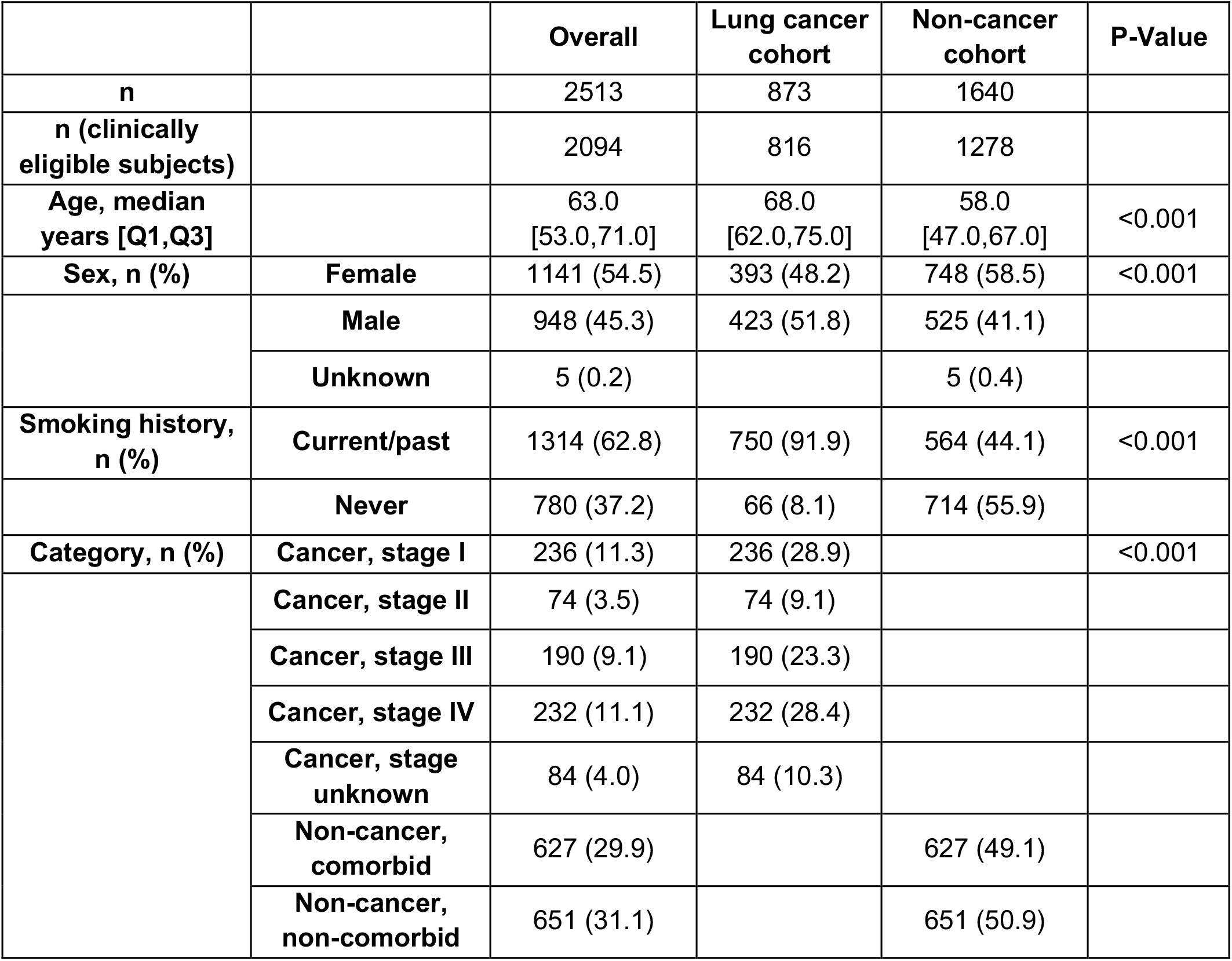
Subject composition in the MOSAIC study.

|  |  | **Overall** | **Lung cancer cohort** | **Non-cancer cohort** | **P-Value** |
| --- | --- | --- | --- | --- | --- |
| **n** |  | 2513 | 873 | 1640 |  |
| **n (clinically eligible subjects)** |  | 2094 | 816 | 1278 |  |
| **Age, median years [Q1,Q3]** |  | 63.0<br>[53.0,71.0] | 68.0<br>[62.0,75.0] | 58.0<br>[47.0,67.0] | <0.001 |
| **Sex, n (%)** | **Female** | 1141 (54.5) | 393 (48.2) | 748 (58.5) | <0.001 |
|  | **Male** | 948 (45.3) | 423 (51.8) | 525 (41.1) |  |
|  | **Unknown** | 5 (0.2) |  | 5 (0.4) |  |
| **Smoking history, n (%)** | **Current/past** | 1314 (62.8) | 750 (91.9) | 564 (44.1) | <0.001 |
|  | **Never** | 780 (37.2) | 66 (8.1) | 714 (55.9) |  |
| **Category, n (%)** | **Cancer, stage I** | 236 (11.3) | 236 (28.9) |  | <0.001 |
|  | **Cancer, stage II** | 74 (3.5) | 74 (9.1) |  |  |
|  | **Cancer, stage III** | 190 (9.1) | 190 (23.3) |  |  |
|  | **Cancer, stage IV** | 232 (11.1) | 232 (28.4) |  |  |
|  | **Cancer, stage unknown** | 84 (4.0) | 84 (10.3) |  |  |
|  | **Non-cancer, comorbid** | 627 (29.9) |  | 627 (49.1) |  |
|  | **Non-cancer, non-comorbid** | 651 (31.1) |  | 651 (50.9) |  |

### Validation of the multi-omics classifier shows high sensitivity and specificity for detection of early-stage lung cancer

Given the high performance of the multi-omics classifier in the training set, we next assessed the sensitivity and specificity of this classifier on the held-out validation set of 398 study subjects (**Table 1**). First, we fixed the decision threshold of the multi-omics classifier to 87.5% sensitivity across all lung cancer stages from the training set (**Methods**) and then evaluated the performance of the classifier at this threshold (henceforth, “model”) in the validation set. Specificity was 89% (95% CI 84-93) and sensitivity was 89% (95% CI 83-93) across all lung cancer stages (**Figure 4**). Sensitivities for stage I, stage II, and stage III-IV (late-stage) lung cancer were 80% (95% CI 68-88), 88% (95% CI 69-98), and 99% (95% CI 94-100), respectively (**Figure 4**). These values were similar to those observed in cross-validation on the training set using the same model.

**Figure 4.**
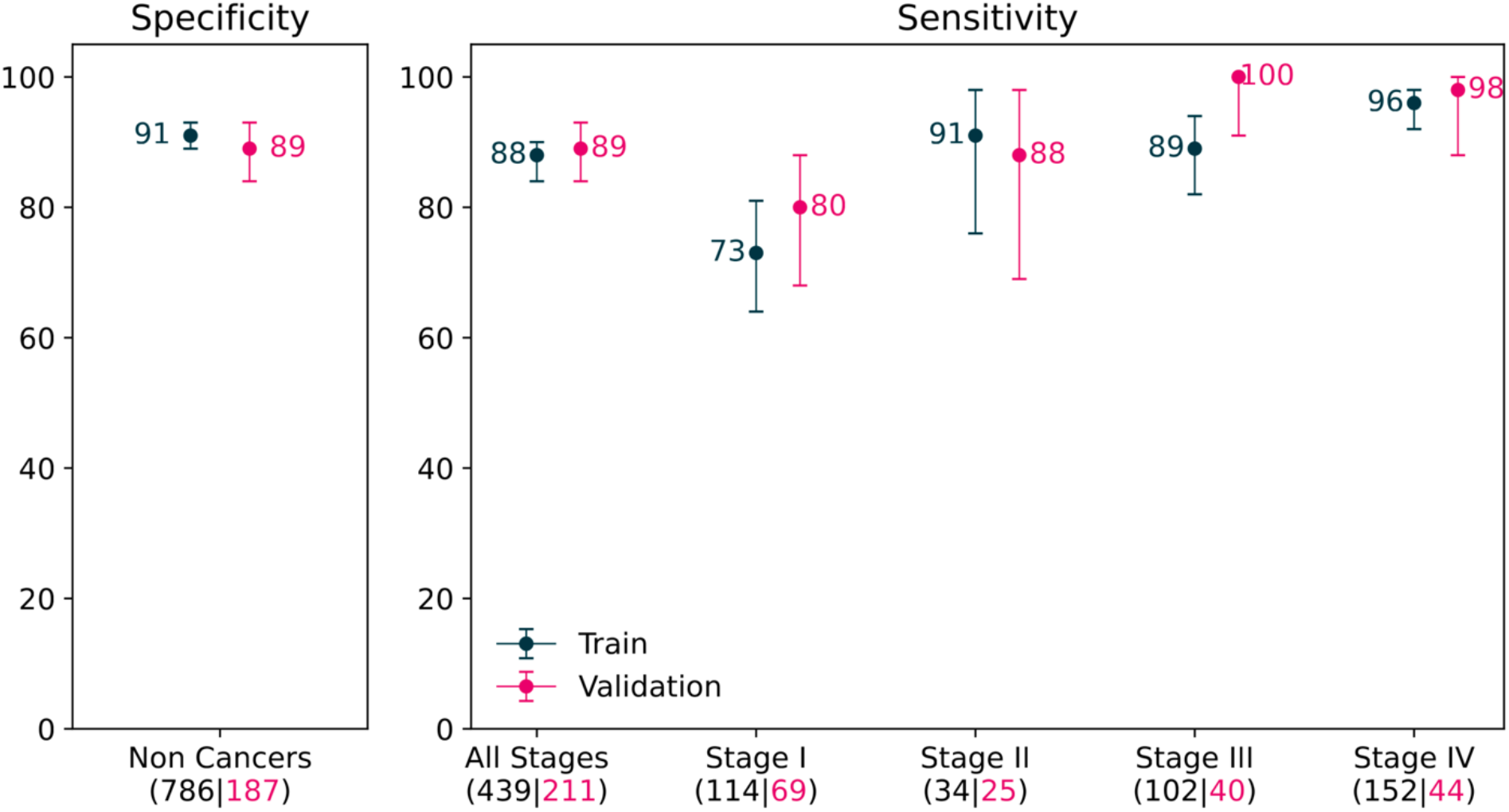
Performance of the validated multi-omics classifier. Stage-wise sensitivity and specificity of the multi-omics classifier for subjects from training (black) and validation (pink) datasets. Error bars indicate 95% Clopper-Pearson confidence intervals. The number of subjects in each sub-group is denoted in parentheses.

### No individual ‘omics type dominate the most important features of the validated model

To gain a broader understanding of the relative contributions of the different ‘omics types to the validated model, the 682 analyte features that comprise the model were ranked based on the mean-information-gain criterion (see **Figure 5** for the ‘omics-type distribution among the top 50 features). 211 of these features were peptide sequences that mapped to 149 distinct proteins, and 354 of the features were transcripts (gene isoforms as well as introns) that mapped to 346 distinct genes. The remaining 117 features were metabolites from 77 distinct metabolic pathways. No individual ‘omics type appeared over-represented among these features. At least 2 features from each ‘omics type were present in the top 20 features, further underscoring the complementary information coming from the different ‘omics types and the importance of a multi-omics approach to enhance classifier performance.

**Figure 5.**
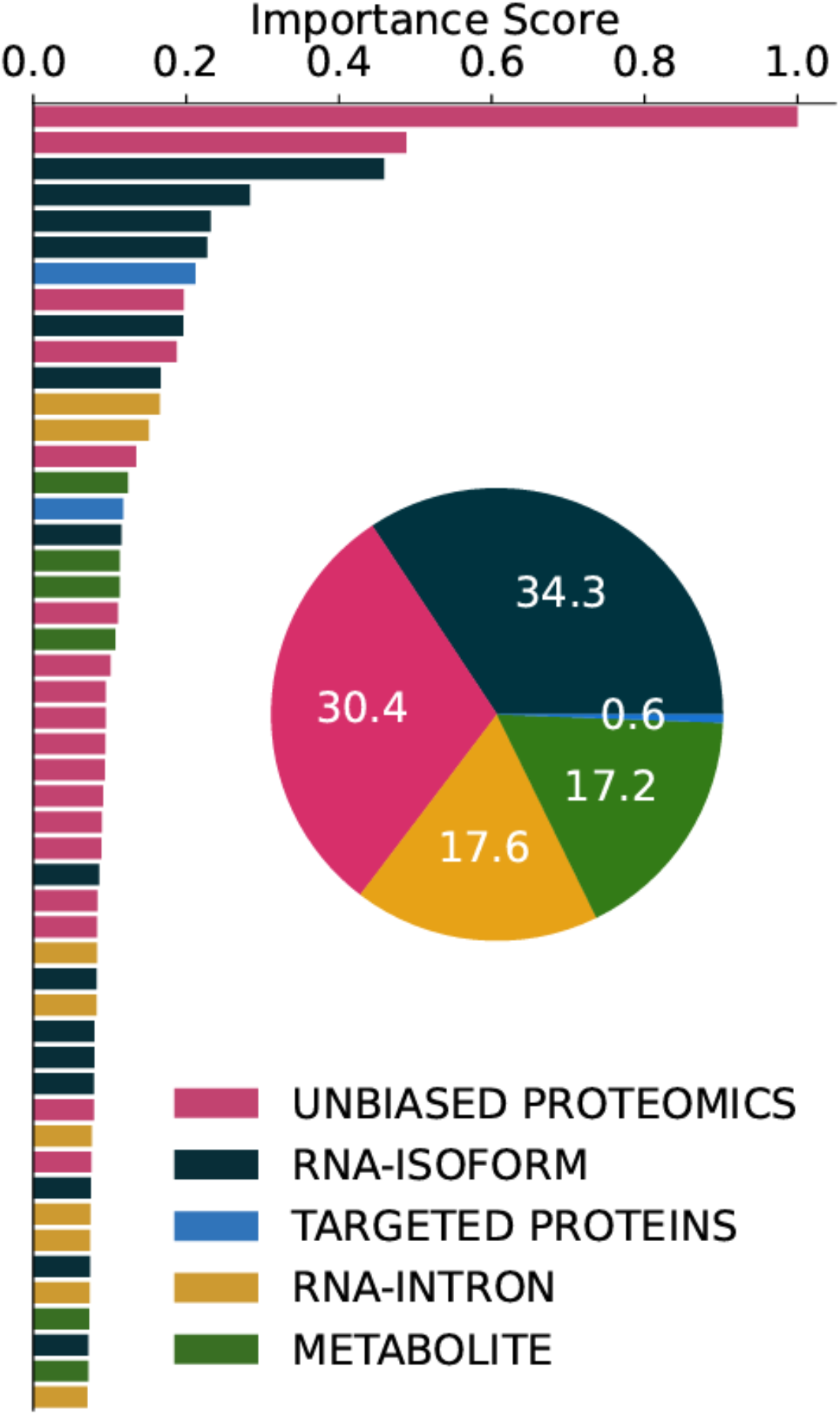
**Features of the multi-omics model ranked by importance score as determined by information gain**. Colors indicate ‘omics type. For ease of interpretation, only importance scores for the top 50 features are shown.

### The important features of the validated model associate with cancer stage progression

To investigate the biological significance of the 682 analyte features of the validated model, we evaluated if the abundance of each individual feature trended with lung cancer stage. Of the 682 features, 412 (60.4%) were significantly associated with cancer stage (Bonferroni-corrected p-value ≤ 0.05; **Figure 6**). This finding suggests that individual features of the validated model might themselves be informative of cancer pathophysiology.

**Figure 6.**
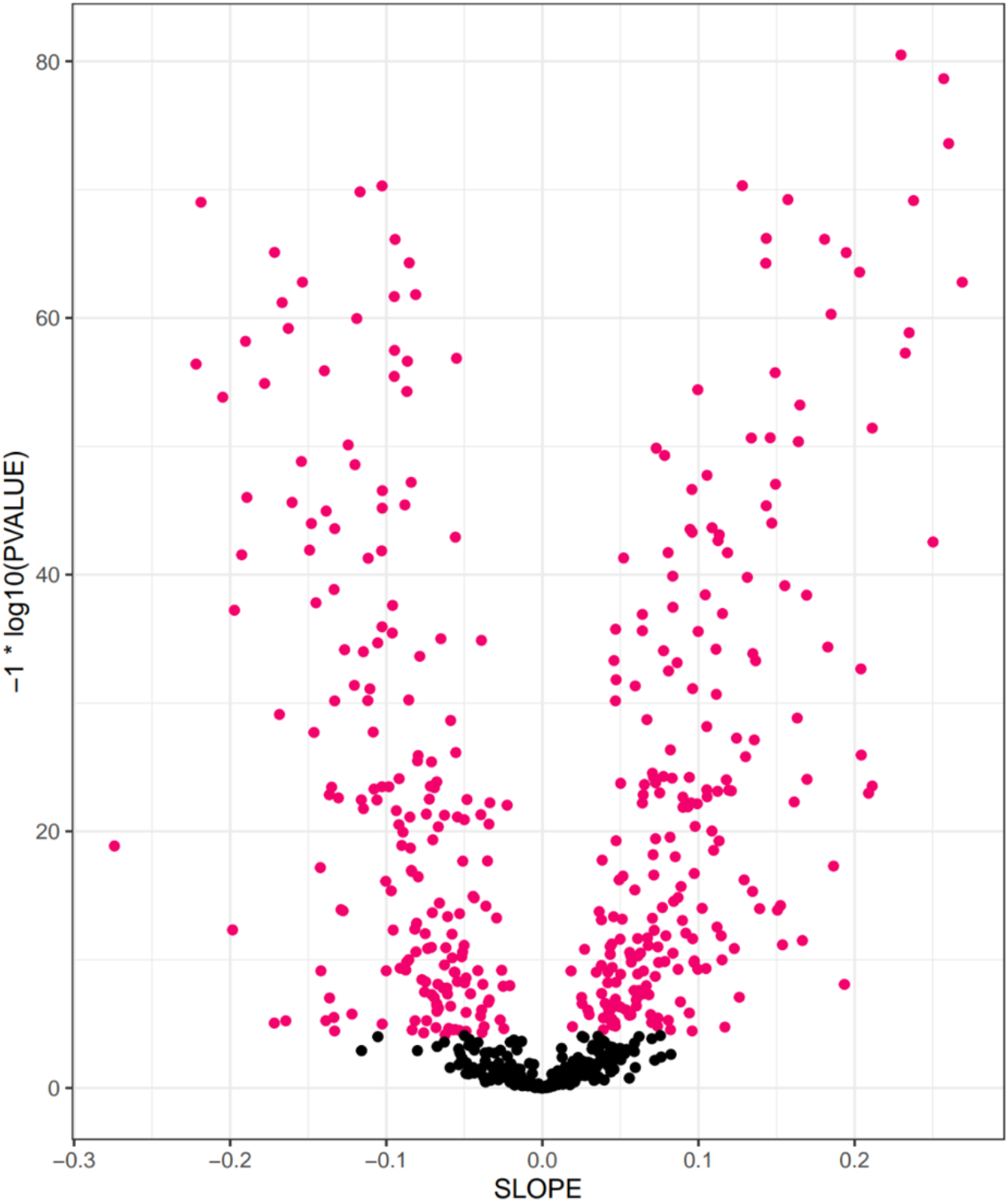
Association of analyte feature abundance and lung cancer stage. Plot of the slope and statistical significance of all 682 features from the validated multi-omics classifier with respect to association with cancer stage. Features with a statistically significant association to cancer stage (Bonferroni-corrected p-value ≤ 0.05) are denoted in pink.

## Discussion

This study represents the largest deep multi-omics interrogation to date, with a lung cancer case-control cohort of 2513 subjects. We leveraged an untargeted approach to collect and analyze over 750,000 orthogonal protein, transcript, and metabolite features that were used to train a machine learning-based lung cancer classifier. This multi-omics classifier detected all-stage and early-stage lung cancer with high specificity and sensitivity in a held-out validation set. The unprecedented breadth and depth of this multi-omics approach presents new opportunities for biomarker discovery. This variety of newly accessible blood analytes has not been adequately represented in previous individual-or multi-analyte approaches that focus on capturing disease signals wholly from genetic material. As the features of the validated model comprise a mix of proteins, transcripts, and metabolites, we hypothesize that different aspects of disease are captured by different ‘omics approaches, suggesting differential and complementary sampling of biological space.

Despite the very large number of individual features detected across all ‘omics types in this study, after modeling, the final multi-omics classifier was composed of a manageable number of features representing 149 distinct proteins, 346 distinct genes, and 77 distinct metabolic pathways that facilitate further development of a practicable assay for early detection of lung cancer. Clinical development of this assay would address a critical clinical need for early cancer detection given the classifier’s performance at detecting stage I disease, which comprised a majority (51.8%) of lung cancer cases in the National Lung Screening Trial.^22^

Although the high performance of the multi-omics classifier suggests a potential future impact on lung cancer health outcomes, there are limitations associated with this study. In particular, although we have designed this study to include all stages of lung cancer, as well as control subjects with pulmonary comorbidities and subjects who smoke, the findings from this work will need to be validated in an appropriately powered prospective study of the intended use population of high-risk individuals aligned with USPSTF, ACS, and NCCN recommendations for annual LDCT screening for lung cancer.^10–12^ This work is ongoing. Despite these caveats, the depth and breadth of the novel biological space interrogated in this study suggest favorable generalization to the prospective intent-to-test setting and supports the further development of a test for the early detection of all-and early-stage lung cancer using plasma from peripheral blood samples.

We have demonstrated the high performance of a novel, untargeted multi-omics biomarker discovery approach with unprecedented interrogative depth and breadth for the early detection of all-and early-stage lung cancer. This platform is generally extensible to additional applications, such as companion diagnostics, recurrence monitoring, and minimal residual disease testing.^49^ Given the potential broad clinical utility of the multi-omics approach demonstrated in this study, we anticipate that the growing number of population studies that collect peripheral blood samples^50,51^ will enable commensurate expansion of multi-omic interrogations of additional complex diseases with great unmet medical need.

## Methods

### Study overview and enrollment criteria

This report describes the MOSAIC study, an observational case-control study of 2513 subjects (**Table 1**) enrolled from 77 unique clinical sites in the US per 2 separate IRB-approved protocols, denoted as 102 and 201. This study was initiated in 2018 and designed to provide peripheral blood samples for discovery and validation of lung cancer biomarkers. All enrolled subjects were adults ≥ 18 years of age and provided written informed consent. Study 102 subjects included diagnosis-aware but treatment-naïve histopathologically confirmed lung cancer subjects (lung cancer cohort) as well as subjects with no prior history of malignancy except non-melanoma skin cancer (non-cancer cohort). Comorbidities of interest, including clinically significant pulmonary (e.g., chronic obstructive pulmonary disease, emphysema, pulmonary fibrosis) and gastrointestinal (e.g., inflammatory bowel disease, pancreatitis, hereditary gastrointestinal cancer syndromes) comorbidities were recorded for all subjects.

Study 201 included subjects without any prior history of malignancy with 1 or more pulmonary nodules that were 6-30 mm in largest diameter and confirmed radiologically prior to enrollment with planned subsequent histopathological characterization. Study 201 subjects were included in both the lung cancer and non-cancer cohorts consistent with histopathological characterization of the biopsied pulmonary nodule(s). A common exclusion criterion for all subjects and studies included the concomitant receipt of biological therapeutics for any indication; no specific small molecule therapeutics for any non-exclusionary condition were prohibited.

Of the 2513 total subjects, 2094 were clinically eligible, consisting of 816 with lung cancers (all stages) and 1278 non-cancer controls (Table 1 and Figure 1B). Malignancies were confirmed histopathologically and staged by the subject’s treating physician(s). At the time of enrollment, subjects with lung cancer were treatment-naïve, with some aware of their diagnosis (Study 102) and others not (Study 201). Pulmonary nodules classified as benign had been either confirmed histopathologically or presumed benign given a history of multiple stable scans over ≥ 1 year consistent with Lung-RADS^®^ 1 guidelines. 276 subjects who had indeterminate pulmonary nodules with nondiagnostic histopathological characterization and/or insufficient radiographic surveillance to support presumptive classification of their pulmonary nodules as benign per stability on successive scans were excluded. Subjects with histopathologically confirmed benign lung pathologies were categorized as non-cancer controls. Non-cancer control subjects with no lung nodules were further categorized as those with and without comorbidities of interest (defined above). Comorbidities were confirmed by subjects’ medical history collected by the participating sites. Of the 1278 non-cancer control subjects, 105 had benign pathologies. Of the remaining 1173 non-cancer subjects, 673 had comorbidities of interest and 500 had neither benign pathologies nor comorbidities of interest.

### Blood sample collection

For all subjects, the median time from enrollment to blood sample collection was 0 days. Per subject, 3 blood samples with a total volume ≤ 50 mL were collected with 3 distinct tube types, specifically dipotassium ethylenediaminetetraacetic acid (K2 EDTA) plasma tubes, serum separator (SST) tubes, and PAXgene^®^ RNA tubes (**Figure 1A**). All sample collection was consistent with manufacturer’s instructions for each tube type. K2 EDTA plasma tubes were centrifuged within 1 hour of collection, and SST tubes were held at room temperature for at least 30 minutes prior to centrifugation. Plasma and serum were aspirated and frozen within 1 hour of centrifugation. Samples were stored at –20°C (or –80°C where available) at the collection site for up to 1 week prior to shipment. Plasma samples, serum samples, and PAXgene^®^ tubes were shipped on dry ice. No additional processing of any tube was performed at the collection site.

### Molecular assay sample processing

Prior to any molecular assay sample processing, study subjects were randomly assigned into either a training set or validation (i.e., testing) set such that clinical site separation was maximized between the 2 partitions. Sample processing to isolate and measure analytes from the corresponding collection tubes was done in a blinded fashion for both the training and validation partitions.

Metabolomics and RNA-seq sample processing were conducted by Metabolon Inc. (Morrisville, NC) and Discovery Life Sciences (Huntsville, AL), respectively. Proteograph™ sample processing and liquid chromatography-mass spectrometry (LC-MS) proteomics data acquisition was done internally and described as follows.

### Proteomics sample processing

A total of 2094 K2 EDTA plasma samples were processed with the Proteograph Assay (Seer, Redwood City, CA) using a 5 nanoparticle (NP1-5) panel following the manufacturer’s protocol. Process control samples were collected, processed, and aliquoted by BioIVT (Westbury, NY). Each batch was balanced to have proportionate representation of cases and controls as well as clinical variables of age, sex, and smoking status. Prior to loading onto the Proteograph instrument, plasma samples were thawed for 60 minutes at 4°C and transferred to 2 mL tubes provided with the Proteograph assay kit. Following Proteograph-processing, the eluted peptide concentration was measured using a quantitative fluorometric peptide assay kit (Cat. No. 23290, Thermo Scientific, Waltham, MA). Following peptide quantification, plates of eluted peptides were dried down in a CentriVap^®^ vacuum concentrator (LabConco, Kansas City, MO) at room temperature overnight and then stored at –80°C. Prior to use, the dried peptide plates were equilibrated at room temperature for 30 minutes and then reconstituted to a concentration of 30 µg/mL for NP1-3,5 and 15 µg/mL for NP4 in a reconstitution buffer (0.1% formic acid [Thermo Fisher, Waltham, MA] in LC-MS grade water [Honeywell, Charlotte, NC] spiked with heavy isotope-labeled retention time peptide standards [iRT, Biogynosys, Switzerland and PepCal, SciEX, Redwood City, CA] prepared according to manufacturer’s instructions). Peptides were fully reconstituted by shaking for 10 minutes at 1000 rpm at room temperature on an orbital shaker and spun down briefly (approximately 10 seconds) in a centrifuge and then loaded onto Evotip separation tips (Evosep, Denmark) following the manufacturer’s protocol. The processed tips were placed on the Evosep One LC system (Evosep, Denmark) and peptides were separated on a reversed-phase 8 cm, 150 μM, 1.5 μM, 100 Å column packed with C18 resin (Pepsep, Germany) using a 60 samples per day (SPD) LC gradient at 40°C (Sonation column oven, Lab Sweden AB).

### LC-MS data acquisition

The LC-MS platform consisted of 4 Evosep One LC systems coupled to 4 timsTOF HT mass spectrometers (Bruker, Germany) set to data independent acquisition (DIA) with parallel accumulation-serial fragmentation (dia-PASEF®) mode.^52^ Proteograph-processed plasma samples were analyzed simultaneously across the 4 LC-MS platforms. A proportional number of samples from subjects with lung cancer and non-cancer control subjects with and without comorbidities of interest were run on each of the 4 LC-MS platforms. Source capillary voltage was set to 1700 V and 200°C. The first MS scan (MS1) to identify peptide precursors was across 100 – 1700 m/z range and an ion mobility window spanning 1/K0 0.75 – 1.31. Peptide precursors were fragmented using collision energies following a linear step-function ranging between 20 eV – 63 eV. Trapped ion mobility spectrometry cell accumulation time was set at 100 milliseconds and the ramp time at 85 milliseconds. For the second MS scan (MS2), variable m/z and ion mobility windows were selected for fragmentation utilizing a Python package for DIA with automated isolation design (py_diAID).^53^

All raw files were analyzed with DIA-Neural Network (NN) (version 1.8.1).^54^ Trypsin protease cleavage with a maximum of 2 missed cleavages was allowed. Cysteine carbamidomethylation was set as fixed modification, while oxidation of methionine and N-terminal protein acetylation were set as variable modifications. MS1 and MS2 mass tolerances were automatically determined by DIA-NN. A cutoff of 1% peptide precursor false discovery rate was used. For other parameters, default DIA-NN settings were applied. DIA-NN outputs were analyzed and visualized with a Python Jupyter notebook and Python packages, pandas (1.5.1),^55,56^ scipy (1.10.1),^57^ numpy (1.23.5),^58^ seaborn (0.12.2),^59^ and matplotlib (3.5.1).^60^

### Data normalization and transformation

Peptide precursor quantity was summed per NP per detected peptide to yield a total intensity for an NP-sequence pair. Modified peptides with different post-translational modifications were treated as different features when summing. Intensity values were natural log-transformed and then DESeq2 normalization^61^ was separately applied to the data for each respective NP.

### Univariate differential analysis

Wilcoxon tests were performed to identify individual, differentially abundant analytes between subjects with lung cancer and non-cancer subjects in the training set. P-values within each ‘omics type were adjusted for multiple hypotheses testing using the Bonferroni correction and a pre-specified threshold of 0.05 was used to denote statistical significance.

### Machine learning and the lung cancer classifier model

Of the 2094 histopathologically confirmed subjects meeting clinical eligibility, 1623 subjects (1225 in training and 398 in validation) were profiled across all molecular assays and passed QC checks on sample contamination and sample swaps.

To train the machine learning model, only subjects from the training set were used. For training of ‘omics-based classifiers, molecular analytes detected in < 25% of training subjects were excluded. Data on all remaining analytes were collated and any remaining missing values were imputed to the minimum value seen across training samples for each respective ‘omics type. For training the baseline classifier built on clinical variables (age, sex, and smoking status), age values were used as-is while sex (male/female) and smoking status (ever/never) categories were one-hot encoded. No exclusions nor imputations were made. Finally, for training of all classifiers, analyte feature values were standardized to zero mean and unit variance across the training subjects. All reference values used for normalization, imputation, and standardization were recorded.

A regularized, tree-based gradient boosted model (XGBoost)^62^ was fitted to the training data using hyperparameters optimized across 10 repeats of 10-fold cross validation. Specifically, for each repeat, the sample mean AUC of each group of hyperparameters was determined based on 10-fold cross validation. The mean of the sampling distribution of sample mean AUCs across the 10 repeats was then calculated and used as the generalized performance estimate of that hyperparameter group. The hyperparameter group with the highest estimated generalized performance was used to fit the final multi-omics lung cancer classifier model on the full training dataset.

To generate a prediction for each subject, the probability value corresponding to 87.5% sensitivity on the training set subjects (across all cancer stages) was selected as the classification threshold for cancer.

### Validation of the multi-omics lung cancer classifier model

As an important safeguard against information leakage that may impact model generalizability, the machine learning team was blinded to the validation data until after classifier training was completed and the trained cancer classifier model was locked. QC checks (used to disqualify samples for inclusion in validation) were defined with the training dataset only to prevent information leakage.

Data from the held-out validation set of 398 subjects were processed in a similar fashion as the training set; however, the reference values used for normalization, imputation, and standardization were based on what was recorded in the training set rather than calculated anew from the validation set.

The trained and locked multi-omics lung cancer classifier was then applied to each subject in the validation set. Specificities and sensitivities for all subgroup analyses (overall and individual stage) were calculated based on these predictions.

### Trend analysis with cancer stage

Ordinary least squares regression was used to fit a univariate model of lung cancer stage across subjects in the training set to each of the 682 analyte features. Non-cancer subjects were encoded as 0, and subjects with lung cancer but no stage information were excluded from these analyses. For each model, the fitted coefficient (and statistical significance thereof) of the lung cancer stage was used to indicate the direction of association between cancer stage and the corresponding feature. P-values were adjusted for multiple hypotheses testing using the Bonferroni correction and a pre-specified threshold of 0.05 was used to denote statistical significance.

## Data Availability

The data referenced in this study will be made available upon reasonable written request and following submission to all study institutional review boards and approval at all participating sites

## Acknowledgements

Medical writing and editing support were provided by Prescott Medical Communications Group (Chicago, IL).

The authors thank Sangeet Adhikari, Isabella Bonomi, Yuya Kodama, Isaiah Odoyo, Preethi Prasad, Hoi-Ting Quanrud, and Sayee Sawale for their support of the MOSAIC study.

## Financial support

This study was funded in its entirety by PrognomiQ, Inc.

## Conflict of interest disclosure statement for all authors

Authors with PrognomiQ, Inc. affiliation are (or were) employees of PrognomiQ, Inc. at the time of study completion and receive (or received) salary and equity compensation as such.

## Notes

### Competing Interest Statement

All authors are current or former employees and shareholders of PrognomiQ and have declared no other conflicts of interest.

### Funding Statement

This work was supported by PrognomiQ. PrognomiQ was involved in the study design, study execution, analysis and interpretation (no grant number)

### Author Declarations

Western Institutional Review Board (WIRB) of WCG gave ethical approval for this work

